# The first wave of the Spanish COVID-19 epidemic was associated with early introductions and fast spread of a dominating genetic variant

**DOI:** 10.1101/2020.12.21.20248328

**Authors:** Mariana G. López, Álvaro Chiner-Oms, Darío García de Viedma, Paula Ruiz-Rodriguez, Maria Alma Bracho, Irving Cancino-Muñoz, Giuseppe D’Auria, Griselda de Marco, Neris García-González, Galo Adrian Goig, Inmaculada Gómez-Navarro, Santiago Jiménez-Serrano, Llúcia Martinez-Priego, Paula Ruiz-Hueso, Lidia Ruiz-Roldán, Manuela Torres-Puente, Juan Alberola, Eliseo Albert, Maitane Aranzamendi Zaldumbide, María Pilar Bea-Escudero, Jose Antonio Boga, Antoni E. Bordoy, Andrés Canut-Blasco, Ana Carvajal, Gustavo Cilla Eguiluz, Maria Luz Cordón Rodríguez, José J. Costa-Alcalde, María de Toro, Inmaculada de Toro Peinado, Jose Luis del Pozo, Sebastián Duchêne, Jovita Fernández-Pinero, Begoña Fuster Escrivá, Concepción Gimeno Cardona, Verónica González Galán, Nieves Gonzalo Jiménez, Silvia Hernáez Crespo, Marta Herranz, José Antonio Lepe, José Luis López-Hontangas, Maria Ángeles Marcos, Vicente Martín, Elisa Martró, Ana Milagro Beamonte, Milagrosa Montes Ros, Rosario Moreno-Muñoz, David Navarro, José María Navarro-Marí, Anna Not, Antonio Oliver, Begoña Palop-Borrás, Mónica Parra Grande, Irene Pedrosa-Corral, Maria Carmen Perez Gonzalez, Laura Pérez-Lago, Luis Piñeiro Vázquez, Nuria Rabella, Jordi Reina, Antonio Rezusta, Lorena Robles Fonseca, Ángel Rodríguez-Villodres, Sara Sanbonmatsu-Gámez, Jon Sicilia, María Dolores Tirado Balaguer, Ignacio Torres, Alexander Tristancho, José María Marimón, Mireia Coscolla, Fernando González-Candelas, Iñaki Comas, on behalf of the SeqCOVID-SPAIN consortium

## Abstract

The COVID-19 pandemic has shaken the world since the beginning of 2020. Spain is among the European countries with the highest incidence of the disease during the first pandemic wave. We established a multidisciplinar consortium to monitor and study the evolution of the epidemic, with the aim of contributing to decision making and stopping rapid spreading across the country. We present the results for 2170 sequences from the first wave of the SARS-Cov-2 epidemic in Spain and representing 12% of diagnosed cases until 14^th^ March. This effort allows us to document at least 500 initial introductions, between early February-March from multiple international sources. Importantly, we document the early raise of two dominant genetic variants in Spain (Spanish Epidemic Clades), named SEC7 and SEC8, likely amplified by superspreading events. In sharp contrast to other non-Asian countries those two variants were closely related to the initial variants of SARS-CoV-2 described in Asia and represented 40% of the genome sequences analyzed. The two dominant SECs were widely spread across the country compared to other genetic variants with SEC8 reaching a 60% prevalence just before the lockdown. Employing Bayesian phylodynamic analysis, we inferred a reduction in the effective reproductive number of these two SECs from around 2.5 to below 0.5 after the implementation of strict public-health interventions in mid March. The effects of lockdown on the genetic variants of the virus are reflected in the general replacement of preexisting SECs by a new variant at the beginning of the summer season. Our results reveal a significant difference in the genetic makeup of the epidemic in Spain and support the effectiveness of lockdown measures in controlling virus spread even for the most successful genetic variants. Finally, earlier control of SEC7 and particularly SEC8 might have reduced the incidence and impact of COVID-19 in our country.

## MAIN

The new coronavirus disease (COVID-19) caused by SARS-CoV-2 emerged in China in October/November 2019^1^ and by the end of March of 2020 it was present in most countries of the world. The World Health Organization declared the new disease as a pandemic on 11^th^ March 2020. Spain suffered a severe epidemic with the first case notified on 29^th^ January^2^ and with an accumulated number of 261,584 cases by 1^st^ July, including 29,965 fatalities. Furthermore, a nationwide seroprevalence study showed that only one in ten cases of infection by SARS-CoV-2 were diagnosed and declared in that period^3^, suggesting that the total number of infections has been vastly underestimated. Spain ordered a series of non-pharmaceutical intervention measures including a general lockdown on 14^th^ March^4^, later applied by many other countries, and was successful in bending the curve by the end of May. Despite these measures, at least 30,000 individuals died during the first wave of the epidemic and a second wave of COVID-19 slowly started by the end of July 2020^5^.

Despite the high incidence accumulated across the country some regions had significantly higher incidence than others. Genomic epidemiology and phylodynamics^6–8^ offer a unique opportunity to understand the early events of the epidemic at the global, regional and local levels, to track the evolution of the epidemic after its initial stages and to quantify the impact of lockdown measures on the genetic variants of the virus. However, there are challenges and caveats that prevent the use of pathogen genomes as the sole source of interpretation. While there is now a large number of SARS-CoV-2 sequences deposited in the databases^9^ there are still important unsampled areas of the world, including some that played an important role in the initial spread of the epidemic. In addition, the virus spreads faster than it evolves^10,11^ which limits the resolution of phylogenetic and phylodynamic analysis^12^. Finally, despite important efforts by sequencing consortiums, only a fraction of the total number of infections has been sequenced. Nevertheless, genomic epidemiology has played an important role in understanding the global and local epidemiology of COVID-19^13–15^.

After the pandemic was declared in Spain, we assembled the National Consortium of genomic epidemiology of SARS-CoV-2 (http://seqcovid.csic.es/). This established a unique network incorporating more than 50 hospitals and scientific institutions across the country to collect clinical samples and epidemiological information from COVID-19 cases. Here we present the results of this nation-wide effort. We were able to sequence 12% of the reported cases before the national lockdown, and 1% of the reported cases of the first wave (until 14^th^ May), including samples of SARS-CoV-2 across Spain in the early months of the pandemic (February-May). Using a combination of pathogen genomics, phylogenetic tools, clinical and epidemiological data we have been able to dissect the very early events in the dispersion of SARS-CoV-2 throughout Spain as well the evolution of the virus during the exponential phase and after the lockdown. We document simultaneous introductions in the country from multiple sources. We show that up to 40% of cases were caused by two Spanish epidemic clades, named SEC7 and SEC8. Seven other Spanish epidemic clades were detected but their role was minor, probably because they were introduced relatively close to the lockdown and had no opportunities for a rapid exponential expansion as the initial two clades had. In contrast to other European countries these SECs belong to early lineages in the epidemic (A in Pangolin, 19B in NextStrain). We also show that the reproductive number, R_e_, of the most successful Spanish epidemic clades quickly declined after the implementation of lockdown measures and they were completely absent from samples taken in July-September. Our results suggest that the most successful variants were those associated with earlier introductions but also that their success may depend on the synergy between superspreading events and high mobility. These results also show the effectiveness of lockdown measures in controlling the virus spread and eliminating established successful epidemic clusters from circulation.

### SARS-CoV-2 was introduced multiple times from multiple sources

Our dataset consists of 2,170 sequences from Spain, collected under ethical approval, from 25^th^ February to 22^nd^ June, coinciding with the initial phases of the COVID-19 pandemic in the country (Figure 1a). The most populated Spanish regions were sampled, resulting in a dataset with sequences representing 16 of the 17 administrative regions in which the country is divided (Figure 1b). 1,962 out of the 2,170 (90.4%) samples analyzed here have been sequenced by the SeqCOVID consortium, while the remaining 208 have been generated by independent laboratories and downloaded from GISAID^9^ (Table S1). Spain displayed a particular viral population structure with a higher proportion of lineage A sequences compared to other European countries^16^(Figure 1c). Strains from patients in Spain were more closely related with cases sequenced in China and were the most abundant during the first weeks of the Spanish epidemic. They were replaced by lineage B strains (Figure S1), which differ by at least 6-7 substitutions from lineage A and dominated the beginning of the pandemic in most European countries. In addition, we observed an heterogeneous distribution of the SARS-CoV-2 genetic diversity within Spain, both at the regional and local levels. For example, our analysis shows how viral diversity declined with geographic distance from a large urban outdoor like Valencia (see Supplementary Notes).

**Figure 1.**
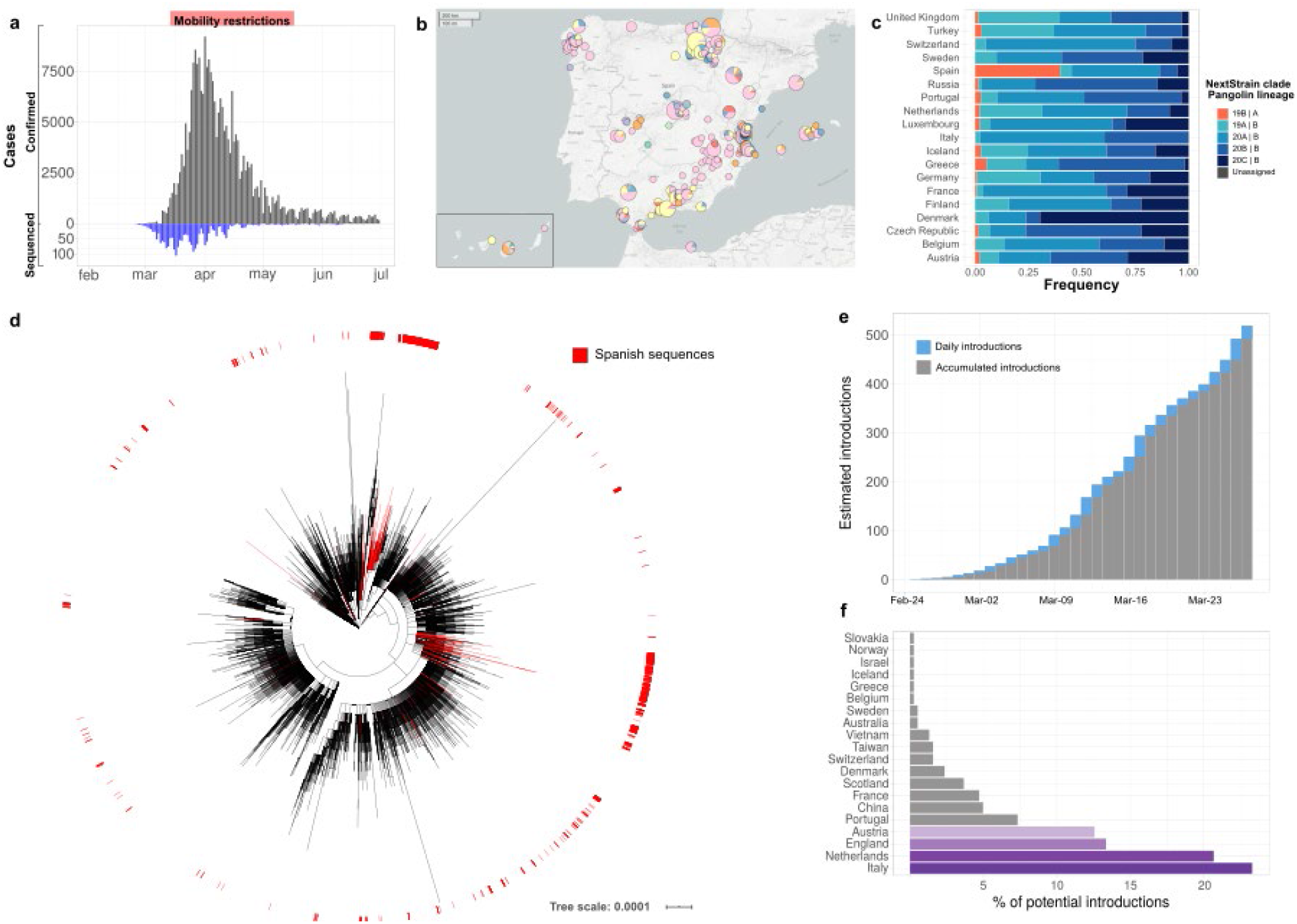
SARS-CoV-2 sequenced genomes from Spain. **a**. Distribution of sequenced samples (blue) versus confirmed cases in Spain (grey) by date. Country lockdown measures were in effect from 13^th^ March to 17^th^ May 2020. **b**. Distribution of the sequenced samples across Spain was plotted in Microreact. This data can be explored with more detail in the Microreact webpage (https://microreact.org/showcase) loading the Data S1 files. The size of each piechart correlates with the number of sequences collected in the corresponding area. Each color corresponds to a specific Pangolin lineage, as detailed in Figure S1 (light yellow and green correspond to lineage A, all the others are lineage B). **c**. Distribution of majorSARS-CoV-2 clades during the first stages of the pandemic (before 1^st^ April 2020), in those European countries with more than 50 sequences deposited in GISAID 13^th^ November 2020. **d**. Global maximum likelihood phylogeny constructed with 32,416 sequences, placement of Spanish samples is indicated in red. **e**. New and accumulated introductions to Spain. Lower-bound introduction estimates were defined as the date of the likely infection of the first case in a cluster (14 days before symptom onset). **f**. Estimated international origin of SARS-CoV-2 introductions based on phylogenetic data; in color, those countries with a likely contribution larger than 10%.

Similarly to other countries^17,18^, phylogenomic analyses suggest the existence of multiple independent entries of the virus into Spain. To identify possible introductions we inspected the placement of Spanish viral samples in a global phylogeny constructed with more than 30,000 sequences (Figure 1d). Given the low genetic diversity of the virus, particularly at the beginning of the epidemic, we found most instances in which a Spanish sample is genetically identical to other variants circulating in the rest of the world. According to their phylogenetic placement, three different possibilities were considered for the phylogenetic position of Spanish sequences. A sequence was included in a ‘candidate transmission cluster’ when it was found in a monophyletic clade with other Spanish sequences; it was included in a ‘zero distance’ group when it grouped with other genetically identical Spanish sequences but also with other foreign sequences; and it was denoted as ‘unique’ when no matching sequence in the Spanish dataset was identified (see detailed definitions of the groups in Mat and Met and in Figure S2). We detected 224 ‘candidate transmission clusters’ comprising 827 sequences (∼40% of the Spanish samples); 30 ‘zero-distance clusters’, comprising 831 sequences, and 513 ‘unique’ sequences (Figures S3). Next, we determined how many unique cases and clusters were compatible with an introduction before the general lockdown. We detected that 191 groups (165 ‘candidate transmission clusters’ plus 26 ‘zero distance clusters’) and 328 unique sequences met this criteria, representing at least 519 independent introductions (distribution of dates in Figure 1e). This is probably an underestimation of the total number of entries because the number of sequences analyzed is a small subset of the total notified cases (Figure 1a). Phylogenetic analysis suggests that the most likely introductions of cases with a clear phylogenetic link (see Methods) came from Italy, the Netherlands, England, and Austria (accounting for ∼23%, ∼20%, ∼13% and 12% of the cases for which a likely country of origin can be inferred, respectively) (Figure 1f). The observation that more than half of the introduction events detected are unique sequences illustrates the heterogeneous outcome after an introduction, as some events resulted in large epidemiological clusters, and others disappeared leaving almost no trace. A clear example is the first described death in Spain for which we have generated a partial sequence and who was infected in Nepal but who did not generate any identifiable secondary cases in our dataset.

### A few genetic variants dominated the first wave in Spain

To identify those introductions that resulted in sustained transmission and therefore epidemiologically successful in the long-term, we scanned the phylogeny for larger clades mainly composed by Spanish samples (see Mat and Met for criteria). We identified 9 Spanish Epidemic Clades (SEC) distributed across the phylogeny, representing 46% of the total Spanish dataset analyzed (995 out of 2,170 samples) (Figure 2a, Figure S4, Figure S5, Table S1, Table S2). We first noticed that only two SECs encompassed 30% and 10% of all Spanish samples (SEC8 and SEC7, respectively). This implies that the introduction of these two specific genetic variants explains a high proportion of the entire epidemic for the first wave in the country. In fact, they were responsible for 44% of the ‘candidate transmission clusters’ identified before the lockdown (Figure 2b). We then estimated the time of introduction in Spain for the 9 SECs using a Bayesian approach (Table S2). As a conservative estimate we considered the time of introduction as any time between the age of the most recent common ancestor of the SEC and the date of the first Spanish sample (Figure 2c). Thus, we assume that the ancestor of the SEC was not necessarily in Spain.

**Figure 2.**
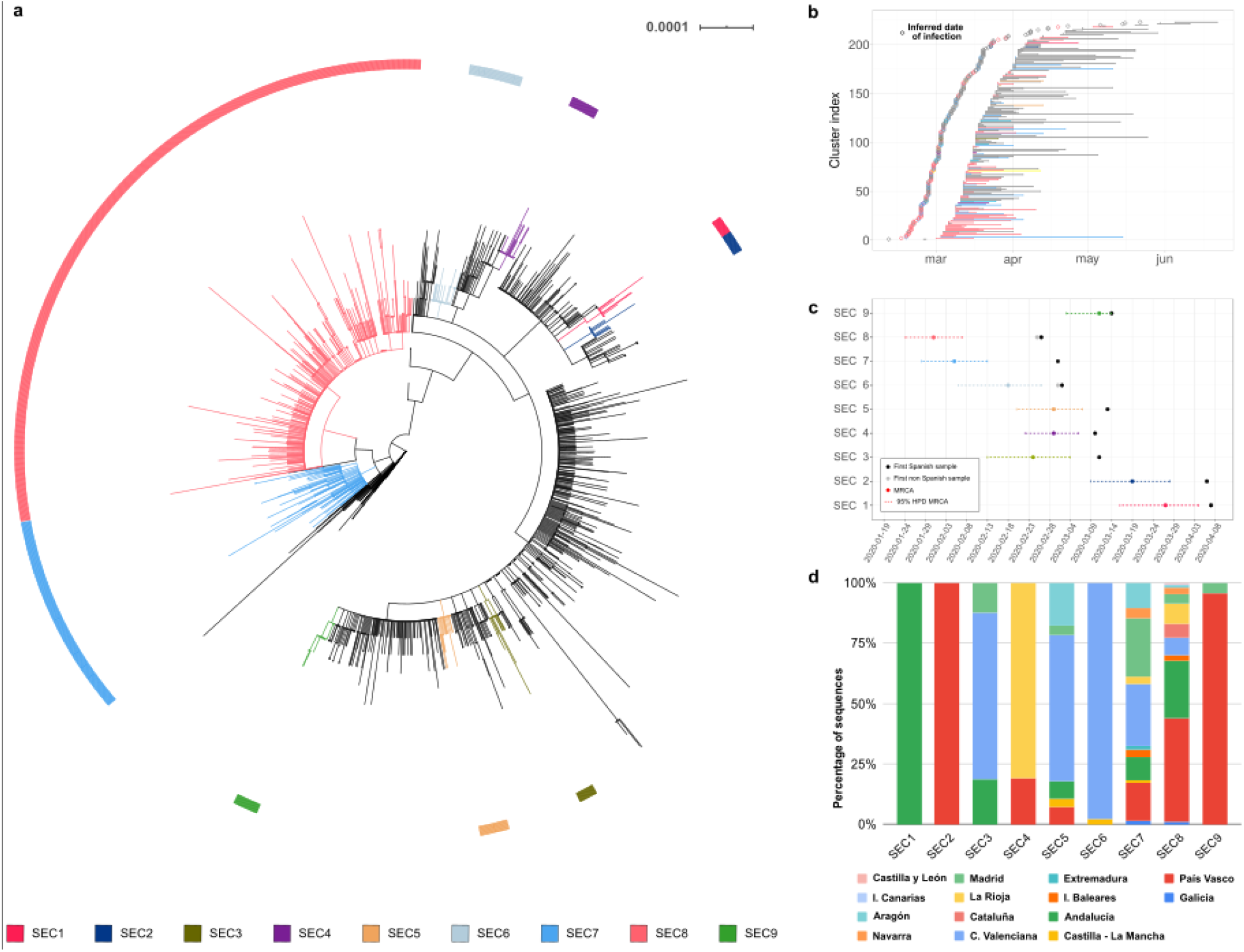
Inferred introduction times and expansion of SECs. **a**. Maximum likelihood phylogenetic tree of Spanish sequences indicating the identified Spanish Epidemic Clades (SECs). **b**. Range of dates for each ‘candidate transmission cluster’ identified within the SECs, and the most probable origin date (14 days before the first documented case) **c**. Time of the Most Recent Common Ancestors (MRCA) of each SEC is plotted, including the 95% HPD interval (High Posterior Density). First collected sample is indicated and pointed if it is Spanish or not. **d**. SEC dispersion through the different regions of the country. Some SECs are circumscribed to one or two regions, while some others have expanded through the complete territory.

Our analysis shows that the earlier the introduction, the larger the size of the SEC (Figure S6). The larger clades, SEC7 and SEC8, were the first successful genetic variants introduced into Spain during late January - February (Figure 2b). Both belong to lineage A (Pangolin nomenclature) and partially explain the peculiar population structure in Spain relative to other European countries (Figure 1c). In addition, compared with other SECs, SEC7 and SEC8 were widely spread in the country, being present in at least 10 of the 17 administrative regions (Figure 2b) and had a mean pairwise geographic distance between samples of more than 300 km regardless whether or not the Islas Canarias and Baleares are included (Figure S7). On the contrary, SECs that were introduced later were smaller and showed a narrower geographic spread (between 0 - 58 km, ANOVA adjusted p-value << 0.01, Supplementary Notes).

### Superspreading events and mobility were key for the success of SEC8

Why some genetic variants succeed over others cannot be answered solely from genomic sequence data. We must also take into account the epidemiological dynamics in the country. There is data supporting a role of the 614G mutation in the spike protein associated with epidemiological success. However, SEC7 and SEC8 do not harbour the variant, explaining why 614G was less frequent during the first weeks of the epidemic in Spain than in other countries (Figue S8). In addition, the inspection of signature positions for both SECs did not lead to any likely genomic determinant of epidemiological success (Table S3). Alternatively, we have investigated linked epidemiological data from the earliest cases to shed light on the early success of SEC8.

In a first phase, SEC8 was introduced at least twice from Italy to the city of Valencia (Figure 3a). There is epidemiological evidence that both cases were infected in Italy, as they attended the Atalanta-Valencia Champions League football match on 19^th^ February, and that one of them initiated a transmission chain upon returning to Valencia a few days later. This epidemiological link strongly suggests that the SEC8 genetic variant was imported from Italy. This introduction occurred in agreement with the estimated time of entry of SEC8 into Spain (Table S2). NextStrain tracking tools for viral spatial spread suggests additional SEC8-related early seedings in Madrid, País Vasco, Andalucía, and La Rioja regions (Video S1) which may involve other countries, not necessarily Italy. Given the lack of virus genetic differentiation and scarce epidemiological information there is no certainty on whether they resulted from independent introductions from abroad or from internal migrations of infected persons, although the simultaneous detection in different regions favours the first option. Most of these multiple introductions occurred during the second half of February, a period in which more than 11,000 daily entries of travelers from Italy were recorded.

**Figure 3.**
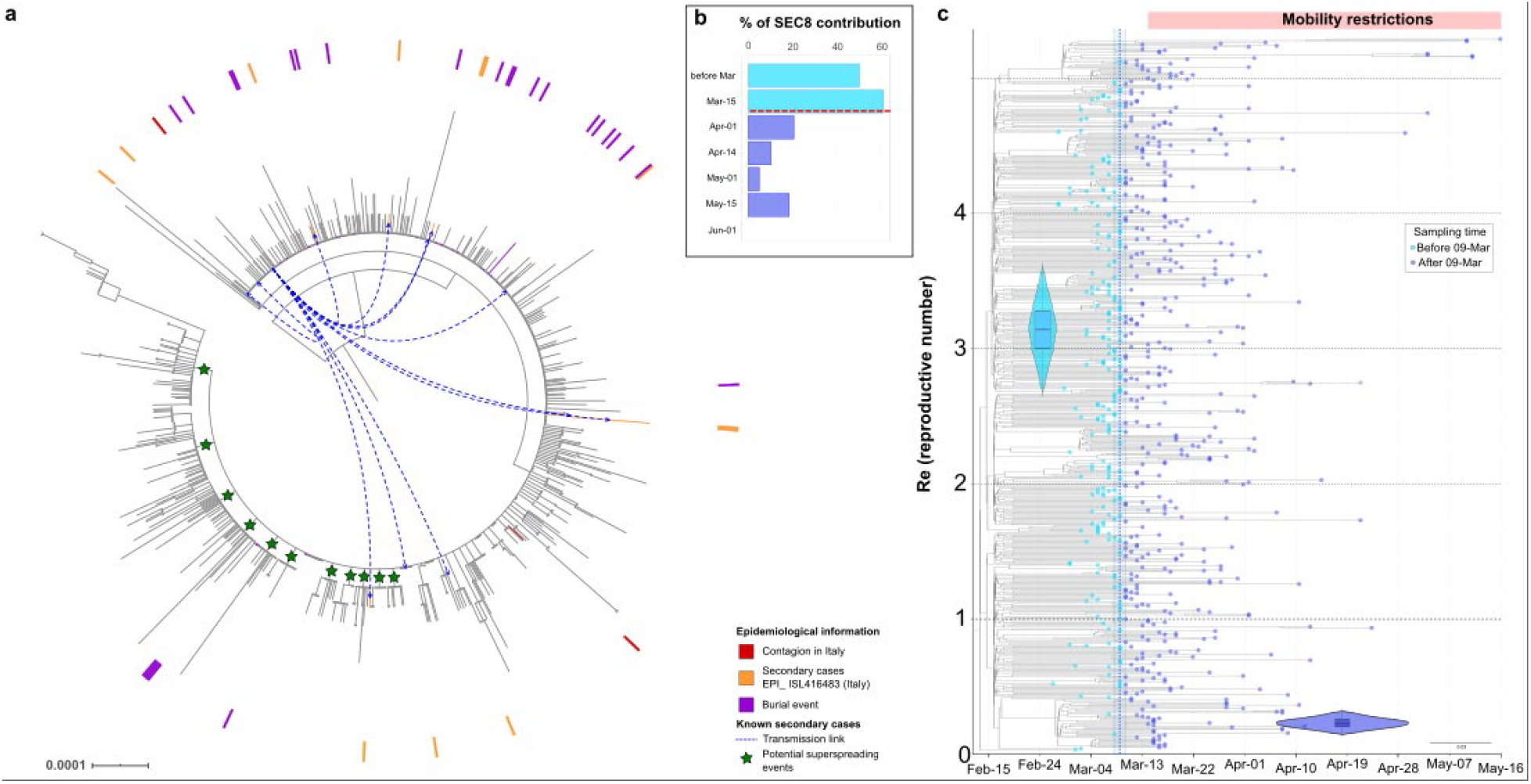
SEC8 epidemiological success and impact of mobility restrictions. **a**. Maximum likelihood phylogeny with all the strains of SEC8. Samples with epidemiological evidence about their origin are marked in the tree. In red, cases imported from different events in Italy. In orange, secondary cases originated from one of the cases introduced from Italy (also marked with blue arrows). In purple, cases related to a large burial in La Rioja. Green stars mark potential superspreading events of more than 10 sequences sharing at least one clade-defining SNP. **b**. Contribution of SEC8 to the total of samples sequenced over time. The horizontal red line marks the start of the Spanish lockdown, on 14^th^ March. **c**. Phylodynamic estimates of the reproductive number (R_e_) of SEC8. The X axis represents time, from the origin of the sampled diversity of SEC8 to the date of the last collected genome on 16^th^ May. The blue dotted line shows the posterior value of the timing of the most significant change in Re, around 9^th^ March [95% HPD: 8–10^th^ March]. The Y axis represents R_e_, and the violin plots show the posterior distribution of this parameter before and after the change time in Re, with a mean of 3.14 [95% HPD: 2.71-3.57] and 0.23 [95% HPD: 0.15-0.32] before and after the change time respectively. The phylogenetic tree in the background is a maximum clade credibility tree with the tips colored according to whether they were sampled before or after 9^th^ March.

In a second phase, SEC8 was fueled by superspreading events. Based on the topology of the phylogenetic tree (Figure 2a) there were multiple clades involving a large number of very closely related sequences (1-3 SNPs) (Figure 3a). Of special relevance was a funeral on 23^rd^ February with attendees from the País Vasco and La Rioja regions from which 25 sequences had been sequenced. Importantly, although they did not differ by more than 2 SNPs these sequences are spread across the SEC8 phylogeny suggesting the existence of many more non-sampled secondary cases across the country (Figure 3a). In a third phase, SEC8, after reaching high frequency locally, was redistributed across the country and in less than two weeks it reached a prevalence of 60% among the sequenced genomes (Figure 3b), being present in almost every region analysed. All these phases occurred between the first known diagnosed SEC8 case on 25^th^ February (Table S2) and the lockdown on 14^th^ March, highlighting the need for very early containment measures to stop the spread of SARS-CoV-2.

### Effect of lockdown on the major clades

In the second half of March, Spain imposed a strict lockdown on non-essential services and movements. A Bayesian birth-death skyline analysis allowed us to evaluate the impact of the lockdown on the effective reproductive number (R_e_) of the most successful SECs. The analyses of SEC7 (Figure S9) and SEC8 (Figure 3c) resulted in similar estimates for R_e_ before the lockdown (2.10 with 95% highest posterior density, HPD:1.67-2.62 and 3.14 HPD: 2.71-3.57, respectively) similar to the R_e_ estimated early in the epidemic for SARS-CoV-2^19,20^. After the lockdown there was a substantial decrease to less than 0.5 in both cases (0.27 95% HPD: 0.06-0.47; 0.23 HPD: 0.15-0.32, respectively). The model also estimated that the date with highest support for a change in R_e_ roughly coincides with the start of the lockdown in Spain on 14^th^ March (20^th^ March HPD: 15-25^th^ March; 9^th^ March HPD: 8-10^th^ March, respectively). In addition, we calculated the doubling time for both SECs^21^. Before the corresponding date of change for R_e_, the doubling time for SEC7 was estimated at 6.3 days (95% HPD: 4.3-10.2 days) and that for SEC8 at 3.3 days (95% HPD: 2.7 - 4.1 days). R_e_ values after those dates had a posterior distribution that did not include 1.0 for both SECs (see Supplementary Notes), a result that supports the reduction in the rate of increase of confirmed cases and that is in agreement with estimates from epidemiological models and data^19,20^.

## DISCUSSION

Our analyses have revealed more than 500 independent introductions of SARS-CoV-2 to Spain between late January, coinciding with the first reported cases in our country^2,22^, and mid-April 2020. The earliest entries corresponded to lineage A, matching the virus diversity profile reported for the country. This lineage was common in Asia but rare in the rest of Europe^23^. We observed that two genetic variants (SEC7 and SEC8) of this lineage dominated the first stages of the epidemic wave in Spain contrary to what was observed in other European countries. In fact, most cases described in Europe at the beginning were lineage B what makes the situation in Spain more unique. This highlights the importance of epidemiological data in which we know that SEC8 was introduced at least from Italy contradicting the dominant lineages in the country at that time^16,24,25.^.

Reasons for why some variants dominate over others can be related to the viral genetics, to founder events associated to particular variants, and to the implementation of different measures over time, not necessarily in an exclusive manner. This variant distribution could also be partly explained by sampling bias. No mutation likely associated with epidemiological success has been identified in our analyses of SEC7 and SEC8 (Table S3). In fact, neither SEC7 nor SEC8 carry the 614G mutation in the spike protein contrary to what is seen in most, but not all, lineage B variants (Figure S8). The mutation 614G has been associated with increased viral shedding compared to the ancestral 614D variant in laboratory conditions^26^ and in transmission studies^27,28^. However, other studies cast doubts on its actual role in the epidemic^29^ suggesting that its impact on epidemic transmission was minor, if any. In the case of Spain, 614G was not behind the initial success of the epidemic because SEC7 and particularly SEC8 were much more common than other genetic variants until the lockdown (10% and 30% of cases respectively). On the contrary, founder events seem to have played an important role for these particular variants. Our analysis shows that they were the first variants introduced in the country and, at least SEC8, were linked to very early superspreading events that contributed to their success. However, an early introduction of lineage A variants also occurred in other European countries but they did not take hold and were displaced by lineage B. Despite the early adoption of strong NPI measures, we hypothesize that epidemic control in the first wave in Spain was soon overwhelmed as compared to countries that controlled early outbreaks^13^. This was likely associated with a strict implementation of the case definition by the WHO, which allowed a stealth dispersion of the first introductions, but also to several superspreading events, which strongly favoured the establishment of the earliest variants arriving into the country. Spain implemented one of the most strict lockdowns in Europe with a high compliance from the population as tracked by mobility data^30^. The efficacy of NPI measures was evident a few weeks later and it was reflected in the almost complete elimination of SEC7 and SEC8 by the end of the first wave. The spread of new variants, represented by other SECs and more isolated cases, corresponded to a new phase in the epidemic at the national level, with much more limited mobility and social interactions which prevented the establishment of large clusters and transmission chains except in high risk settings such as nursing-homes and long-term care facilities.

This study has several limitations. Despite being one of the countries with more contribution to public repositories, our dataset only represents a small subset of confirmed cases that occurred in the first COVID-19 wave (1% of cases). Moreover, sampling across the country was heterogeneous and the representation of each region in the dataset was not always proportional to the incidence during the studied period. Lack of genome data from countries with high disease burden, especially at the beginning of the pandemic, may have led to underestimating the total number of introductions and prevented a reliable identification of their likely sources based only on viral genome sequences. In addition, we did not have access to individual patient data for most cases. Despite these limitations, we have been able to investigate some of the key cases and events that ignited the epidemic in Spain. This allowed us to understand the origin and early spread of SEC8, which would not have been possible based only on genome data. But we have also shown that genetic data can be used to accurately estimate relevant epidemiological parameters such as R_e_ and doubling times even when the proportion of sampling is low.

We believe that our results allow us to draw lessons for the control of this and future pandemics. First, we have shown how specific variants can be used to track the effectiveness of epidemic control measures. In February, the number of SEC8 cases was just a few dozens and yet it ended up accounting for 60% of the sequenced samples in the first weeks of March. Second, the closure of borders to countries with high incidence is relevant to reduce simultaneous and multiple imports of the virus, but its efficacy depends on the inward incidence of the disease^31^. The most successful SECs during the first wave were probably those that arrived early, multiple times, and to diverse locations. Thus, as suggested elsewhere, founder effects are important for the success of certain variants. Third, SEC7 and SEC8 extended across Spain in a matter of days. Controlling mobility is essential when the level of community transmission is high, as demonstrated by the significant decrease in R_e_ for these high-transmission genetic variants after the lockdown. As a comparison, before the lockdown R_e_ values were 50% higher in Spain (3.3 for SEC8) than in Australia (1.63), and they underwent a reduction down to 7% of the original value (0.23) as a result of the containment measures, compared to 30% (0.48) in Australia^15^. From a public health perspective, our results add to the evidence that the success of specific genetic variants is fueled by superspreading events which rapidly increase the prevalence of the virus^32^. Subsequently, coupled to the high mobility of our connected world, a variant may end up dominating the epidemic in a geographic location. This is what occurred to SEC8 and what at a local level has been described in Boston^33^. In fact, we have recently described a new variant in Europe that is rapidly growing in several countries, which is also linked to initial superspreading events^34^. The conclusion is that early diagnosis and notification of cases would have helped to a timely implementation of effective contact tracing that, coupled with earlier mobility closures and maybe tighter border control, would have probably delayed a few days the expansion of genetic variants such SEC8 during the early stages of the epidemic in Spain. Whether this might have changed the global shape of the epidemic in the country or other genetic variants would have performed its role leading to a similar outcome cannot be ascertained, but the comparison with other countries lead us to suspect that there would have been not many differences with them.

## Data Availability

The analysis pipeline used to map and analyze the sequences is available at
https://gitlab.com/fisabio-ngs/sars-cov2-mapping. All the genomic sequences used in the
analyses are available in the GISAID database, and the accession numbers can be found in
Table S1.

## SUPPLEMENTARY MATERIAL

- Supplementary Notes
- Supplementary Material and Methods
- Supplementary References
- Supplementary Data
  - Table S1: GISAID accession numbers for the 32914 sequences used in this study. The ‘basal group’ used for dating and the sequences representative of the pangolin lineages are marked for identification.
  - Table S2: SEC characteristics and inferred origin time. The time of the most recent common ancestors (MRCA) of each SEC was estimated with a Bayesian molecular clock analysis. “MRCA date” indicates the median value for the age of the closest SEC MRCA. The 95% Highest Posterior Density (HPD) credibility interval for this value is provided. “SEC size” indicates the number of samples belonging to each SEC. The first Spanish collected sample within each SEC is also indicated; the inferred date of infection is inferred as the time span between the oldest MRCA date and the first Spanish collected sample. Number of “candidate transmission clusters”, “zero distance clusters” and “unique” included in each SEC are mentioned. “MRCA2” indicates the time of the previous ancestor to the MRCA, considering only nodes that display a posterior probability higher than 0.5. If we consider that the MRCAs were already in Spain, then the introductions into the country occurred between the MRCA2 and the MRCA dates.
  - Table S3: SEC definitory mutations.
  - Data S1.zip: Alignment and phylogenetic tree of the Spanish sequenced samples, to be plotted using the Microreact server.
  - Video S1: Video showing the transmission dynamics of SEC8 within Spain from 2020-01-16 to 2020-07-19, obtained from NextStrain.
- Supplementary Figures
  - Figure S1: Abundance of the different Pangolin lineages in the dataset by epidemiological week (number of weeks since 2019-12-23) as plotted in Microreact.
  - Figure S2: Examples of the different groups of sequences identified. ‘Candidate transmission clusters’ are groups of Spanish sequences that form a clade. ‘Zero distance clusters’ are groups of Spanish sequences that are at zero distance from each other. Finally, the ‘unique’ sequences are Spanish sequences that are more than 1 SNP away from any other Spanish sequence and that do not share a most recent common ancestor (MRCA) node with other Spanish sequences
  - Figure S3: Distribution of the different clusters/groups sizes in Spanish samples.
  - Figure S4: Number of international and Spanish sequences in each SEC.
  - Figure S5: Phylogenetic location of each SEC in the global SARS-CoV-2 phylogeny. Sequences from Spain are coloured according to their SEC (as indicated in Figure 2) while international sequences remain in black colour.
  - Figure S6: Time of the MRCA of each SEC plotted against the contribution of each SEC to the total number of samples in the Spanish dataset. We observed a significant correlation (□=- 0.69, p-value=0.03) between the time of the MRCA of each SEC and its size, estimated as the number of samples sequenced.
  - Figure S7: Distribution of genetic (salmon) versus geographic (grey) distances within each pair of samples belonging to the same SEC.
  - Figure S8: Distribution of sequences harbouring the 614G mutation (blue) versus the 614D mutation (salmon,wild-type) in the S gene for the spanish sequences in our dataset. In the left panel, a histogram of samples sorted by date of sequencing. At right, frequency of both mutations in the sequenced samples by date. The national lockdown event is marked by a purple vertical line.
  - Figure S9: Phylodynamic estimates of the effective reproductive number (R_e_) of Spanish SEC7. A birth–death skyline (BDSKY) model was implemented in Beast v.2, allowing for piecewise changes in Re, with the time and magnitude estimated from the data. The X axis represents time, starting with the MRCA of all sampled diversity within SEC7 and ending with the date of the most recently sequenced genome from 15^th^ May. The blue dotted line indicates the posterior value of the timing of a most significant decrease in Re, around 20^th^ March [95% HPD: 15–25^th^ March]. The Y axis represents Re, and the violin plots show the posterior probability distribution for this parameter before and after the change time in Re; with a mean of 2.10 [95% HPD: 1.67–2.62] and 0.27 [95% HPD:0.06–0.47] before and after this time, respectively. The phylogenetic tree in the background is the maximum clade credibility tree from the BDSKY analysis, with the tips colored according to whether they were sampled before or after 20^th^ March.
  - Figure S10: Mean pairwise genetic distance vs geographic distance (in SNP number), between the largest cities (> 70k inhabitants) of the Comunidad Valenciana autonomous region.
  - Figure S11: Left) Heatmap of genetic diversity for the province of Valencia; red colors indicate high diversity; blue colors indicate lower diversity. Genetic diversity has been measured as the number of base substitutions per site averaged over all sequence pairs within each municipality. Genetic diversity is largest near Valencia, the region’s capital, and decreases with geographic distance to it. Right) All sequences included in our dataset from Comunidad Valenciana, coloured according to the pangolin lineage they belong to.

## Acknowledgements

This work was funded by the Instituto de Salud Carlos III project COV20/00140, Spanish National Research Council project CSIC-COV19-021 and ERC StG 638553 to IC, and BFU2017-89594R to FGC. MC is supported by Ramón y Cajal program from Ministerio de Ciencia and grants RTI2018-094399-A-I00 and SEJI/2019/011.

We gratefully acknowledge Hospital Universitari Vall d’Hebron, Instituto de Salud Carlos III, IrsiCaixa AIDS Research Lab and all the international researchers and institutions that submitted sequenced SARS-CoV-2 genomes to the GISAID’s EpiCov™ Database, as an important part of our analyses have been made possible by the share of their work.

## Author contributions

IC, FGC and MC conceived the work. GAG, GDA and SJS set up the bioinformatics environment and the analysis pipeline. MGL, ACO, PRR and NGG analysed the data. ACO and MGL wrote the first version of the draft. AO, JR, EM, AEB, AN, DGV, LPL, MH, JS, MAM, MT, MPBE, NGJ, GM, LMP, PRH, LRR, MTP, IGN, JFP sequenced genomes. GCE, MMR, LPV, JMM, RMM, MDTB, JAL, VGG, ARV, DN, EA, IT, ACB, SHC, MLCR, AR, AT, AMB, JMNM, IPC, SSG, BFE, CGC, BPB, ITP, AC, VM, MPG, LRF, JLP, JA, JJCA, MCPG, JAB, NR, JLLH, MAZ provided samples. IC, FGC, MC, ACO, MGL, SD and DGV critically reviewed and contributed to the final version of the paper.

## Annex I - List of the SeqCOVID-SPAIN consortium members

Iñaki Comas, Fernando González-Candelas, Galo Adrian Goig, Álvaro Chiner-Oms, Irving Cancino-Muñoz, Mariana Gabriela López, Manoli Torres-Puente, Inmaculada Gómez-Navarro, Santiago Jiménez-Serrano, Lidia Ruiz-Roldán, María Alma Bracho, Neris García-González, Llúcia Martínez-Priego, Inmaculada Galán-Vendrell, Paula Ruiz-Hueso, Griselda De Marco, M^a^ Loreto Ferrús, Sandra Carbó-Ramírez, Mireia Coscollá, Paula Ruiz-Rodríguez, Giuseppe D’Auria, Francisco Javier Roig Sena, Isabel Sanmartín, Daniel García-Souto, Ana Pequeño-Valtierra, Jose M. C. Tubio, Javier Temes, Jorge Rodríguez-Castro, Martín Santamarina, Nuria Rabella, Ferrán Navarro, Elisenda Miró, Manuel Rodríguez-Iglesias, Fátima Galán-Sanchez, Salud Rodríguez-Pallares, María de Toro, María Pilar Bea-Escudero, José Manuel Azcona-Gutiérrez, Miriam Blasco-Alberdi, Alfredo Mayor, Alberto L. García-Basteiro, Gemma Moncunill, Carlota Dobaño, Pau Cisteró, Darío García de Viedma, Laura Pérez-Lago, Marta Herranz, Jon Sicilia, Pilar Catalán, Julia Suárez, Patricia Muñoz, Cristina Muñoz-Cuevas, Guadalupe Rodríguez Rodríguez, Juan Alberola, Jose Miguel Nogueira Coito, Juan José Camarena Miñana, Antonio Rezusta, Alexander Tristancho, Ana Milagro Beamonte, Nieves Martínez Cameo, Yolanda Gracia Grataloup, Elisa Martró, Antoni E. Bordoy, Anna Not, Adrián Antuori, Anabel Fernández, Nona Romaní, Rafael Benito, Sonia Algarate Cajo, Jessica Bueno, Jose Luis del Pozo, Jose Antonio Boga, Cristián Castelló Abietar, Susana Rojo Alba, Marta Elena Álvarez Argüelles, Santiago Melón García, Maitane Aranzamendi Zaldumbide, Andrea Vergara, Miguel J Martínez, Jordi Vila, David Posada, Diana Valverde, Nuria Estévez, Iria Fernández-Silva, Loretta de Chiara, Pilar Gallego, Nair Varela, Rosario Moreno-Muñoz, M^a^ Dolores Tirado Balaguer, Ulises Gómez-Pinedo, Mónica Gozalo Margüello, M^a^ Eliecer Cano García, José Manuel Méndez Legaza, Jesús Rodríguez Lozano, María Siller Ruiz, Daniel Pablo Marcos, Antonio Oliver, Jordi Reina, Carla López-Causapé, Andrés Canut-Blasco, Silvia Hernáez Crespo, María Luz Cordón Rodríguez, M^a^ Concepción Lecaroz Agara, Carmen Gómez González, Amaia Aguirre Quiñonero, José Israel López Mirones, Marina Fernández Torres, M^a^ Rosario Almela Ferrer, José Antonio Lepe, Verónica González Galán, Ángel Rodríguez-Villodres, Nieves Gonzalo Jiménez, Maria Montserrat Ruiz García, Antonio Galiana, Judith Sánchez-Almendro, Gustavo Cilla Eguiluz, Milagrosa Montes Ros, Luis Piñeiro Vázquez, Ane Sorrarain, José María Marimón, Maria Dolores Gómez Ruiz, Eva M. González-Barberá, José Luis López-Hontangas, José María Navarro-Marí, Irene Pedrosa-Corral, Sara Luisa Sanbonmatsu-Gámez, Maria Carmen Perez Gonzalez, Francisco Javier Chamizo López, Ana Bordes Benítez, David Navarro, Eliseo Albert, Ignacio Torres, María Isabel Gascón Ros, Cristina Torregrosa Hetland, Eva Pastor Boix, Paloma Cascales Ramos, Begoña Fuster Escrivá, Concepción Gimeno Cardona, María Dolores Ocete Mochón, Rafael Medina González, Julia González-Cantó, Olalla Martínez, Begoña Palop-Borrás, Inmaculada de Toro Peinado, María Concepción Mediavilla Gradolph, Oscar González-Recio, Mónica Gutiérrez-Rivas, Encarnación Simarro Córdoba, Julia Lozano Serra, Mónica Parra Grande, Lorena Robles Fonseca, Adolfo de Salazar, Laura Viñuela, Natalia Chueca, Federico García, Cristina Gomez-Camarasa, Ana Carvajal, Vicente Martín, Juan Miguel Fregeneda-Grandes, Antonio José Molina, Héctor Argüello, Tania Fernandez-Villa, Amparo Farga Martí, Rocío Falcón, Victoria Domínguez Márquez, José Javier Costa-Alcalde, Rocío Trastoy, Gema Barbeito-Castiñeiras, Amparo Coira, María Luisa Pérez del Molino Bernal, Antonio Aguilera, Anna Planas, Álex Soriano, Israel Fernández-Cádenas, Jordi Pérez-Tur, María Ángeles Marcos, Manuel Segovia Hernández, Antonio Moreno Docón, Juan Carlos Galan, Esther Viedma Moreno, Jesús Mingorance, Jovita Fernández-Pinero, Elisa Rubio García, Aida Peiró-Mestres, Jessica Navero-Castillejos

