## Supplementary figures and images for "The first wave of the Spanish COVID-19 epidemic was associated with early introductions and fast spread of a dominating genetic variant"

### Figure S5.pdf

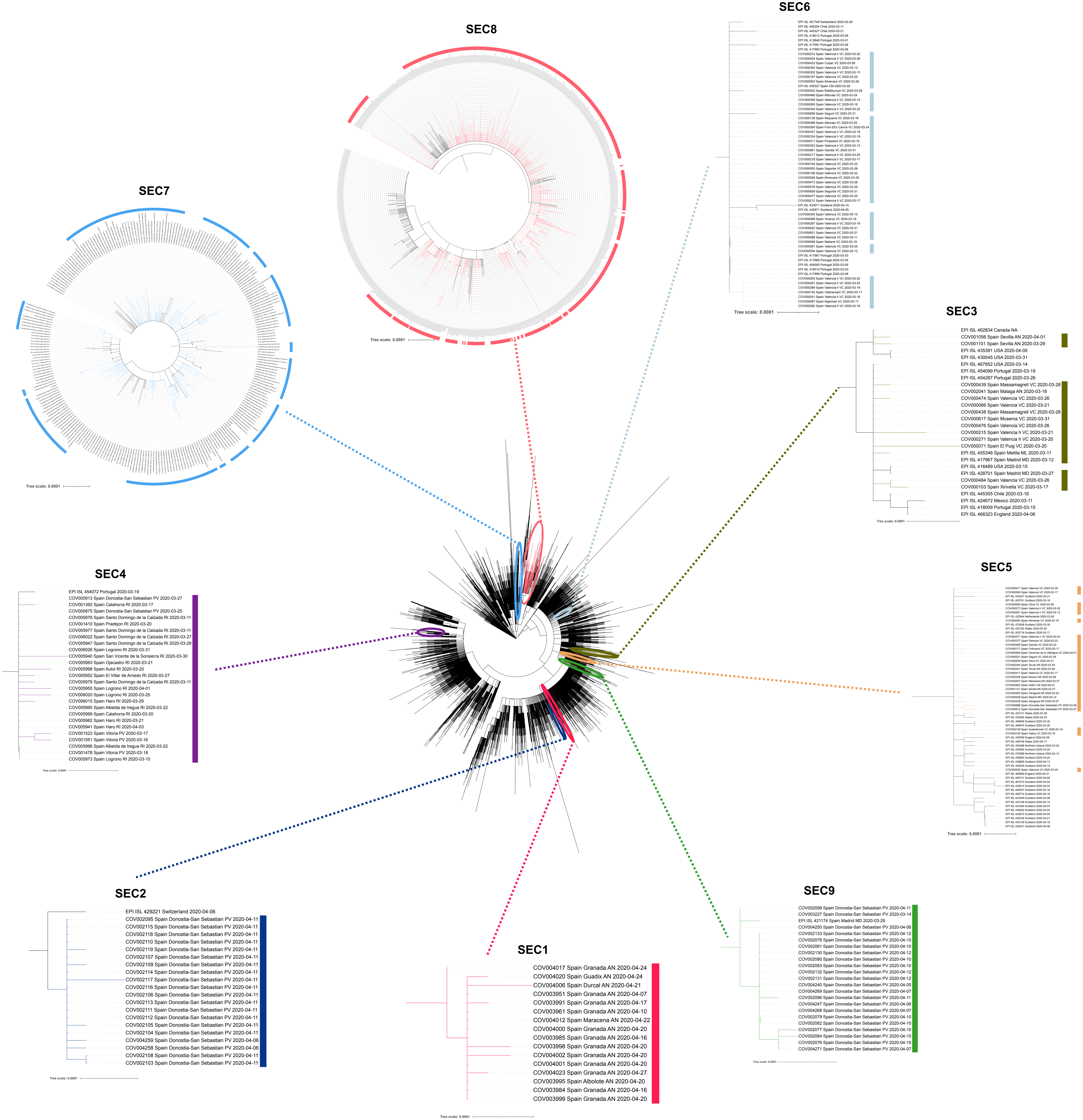

### Figure_S1.png

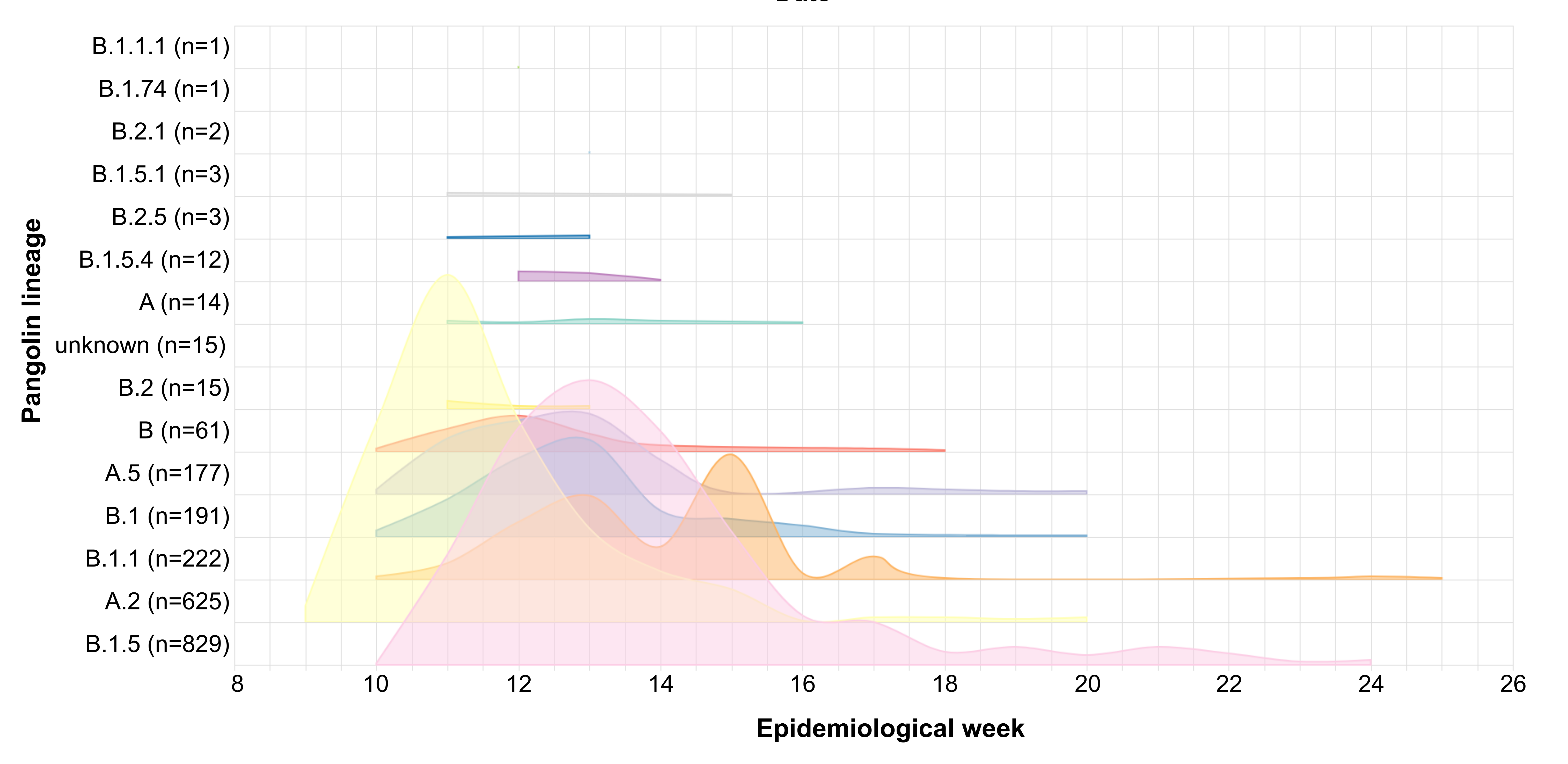

### Figure_S2.png

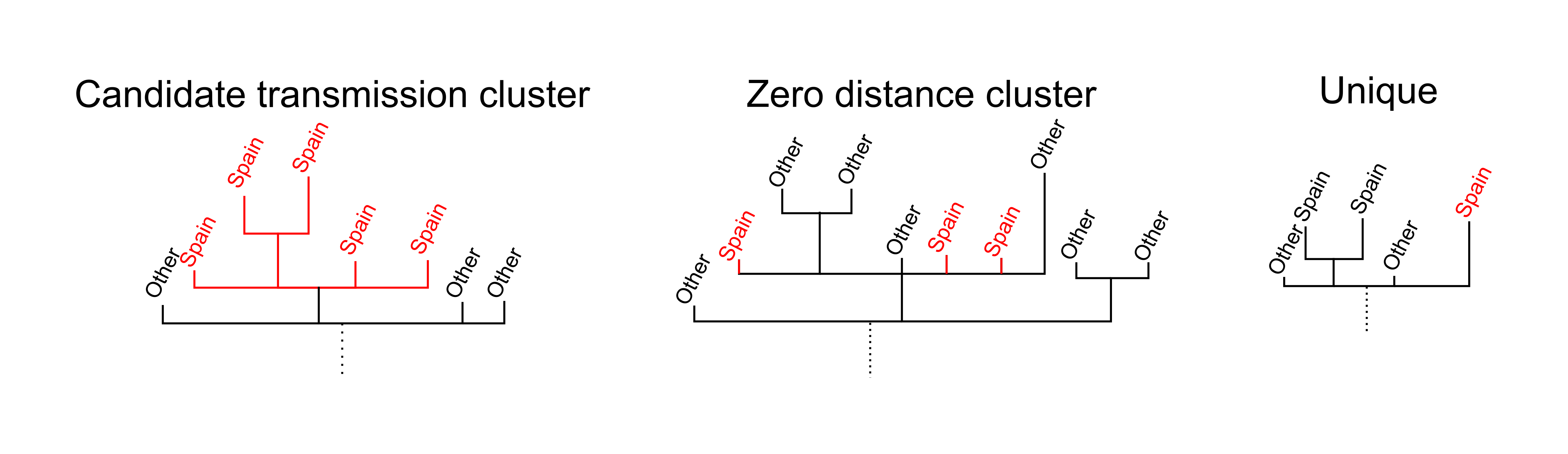

### Figure_S3.png

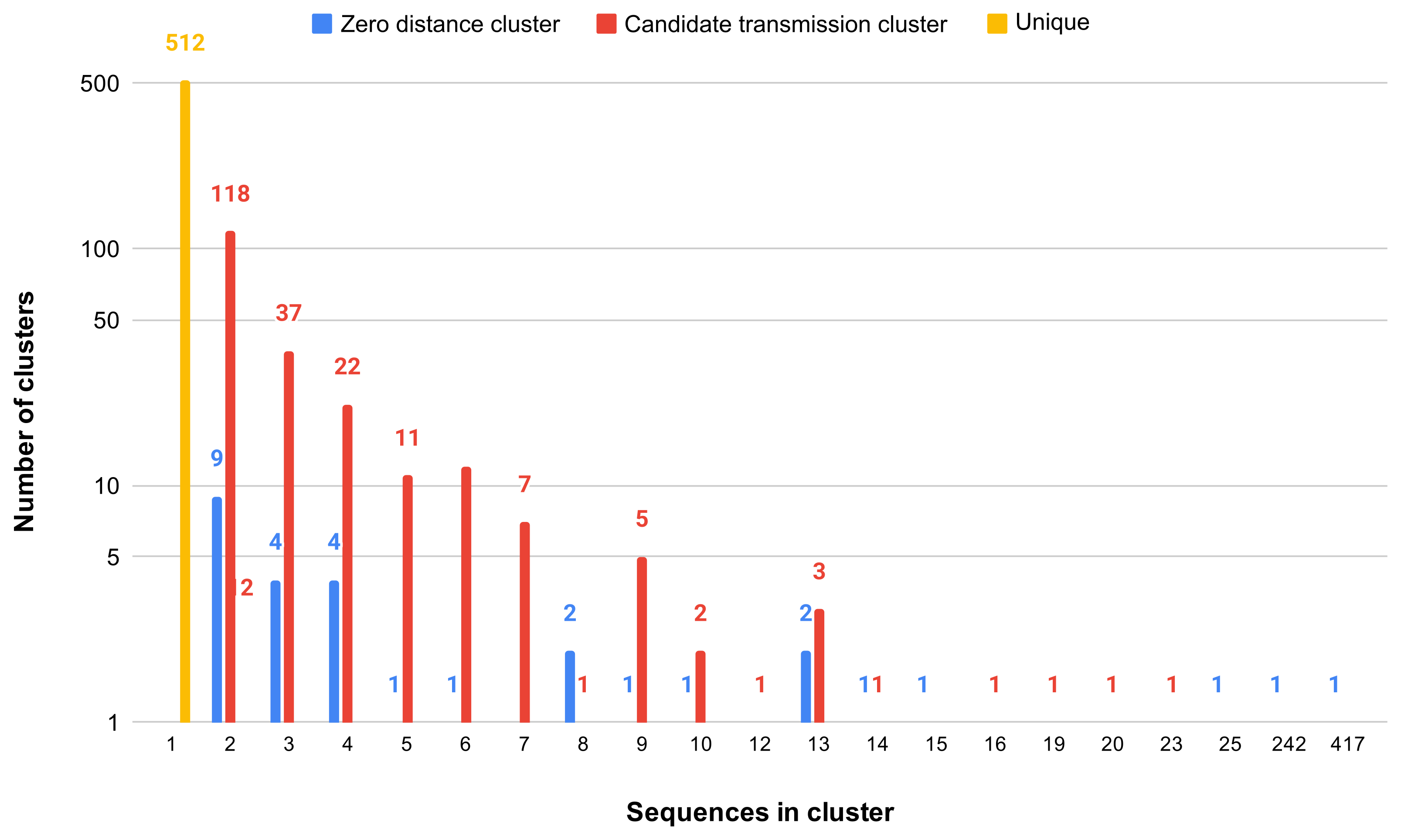

### Figure_S4.png

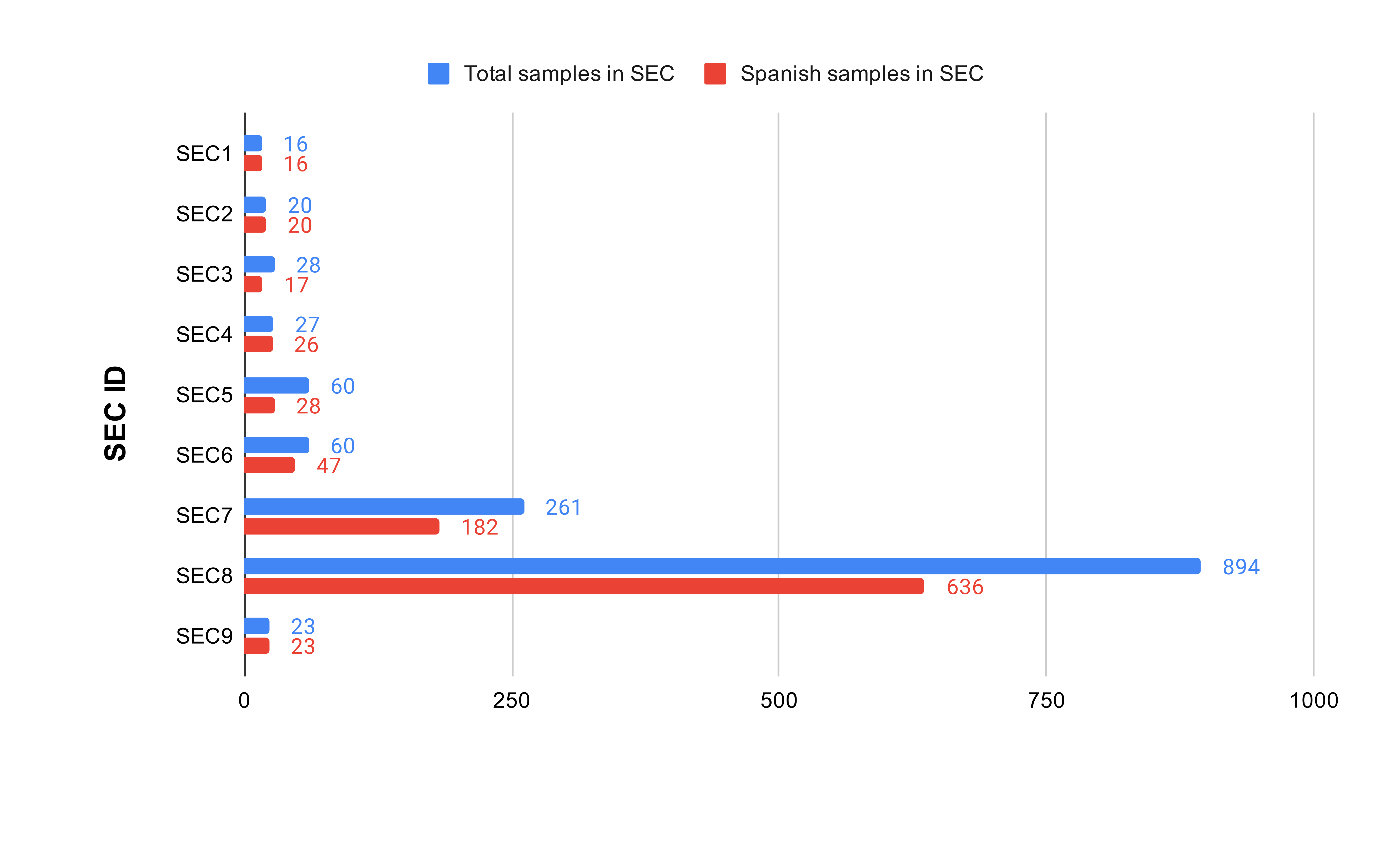

### Figure_S6.png

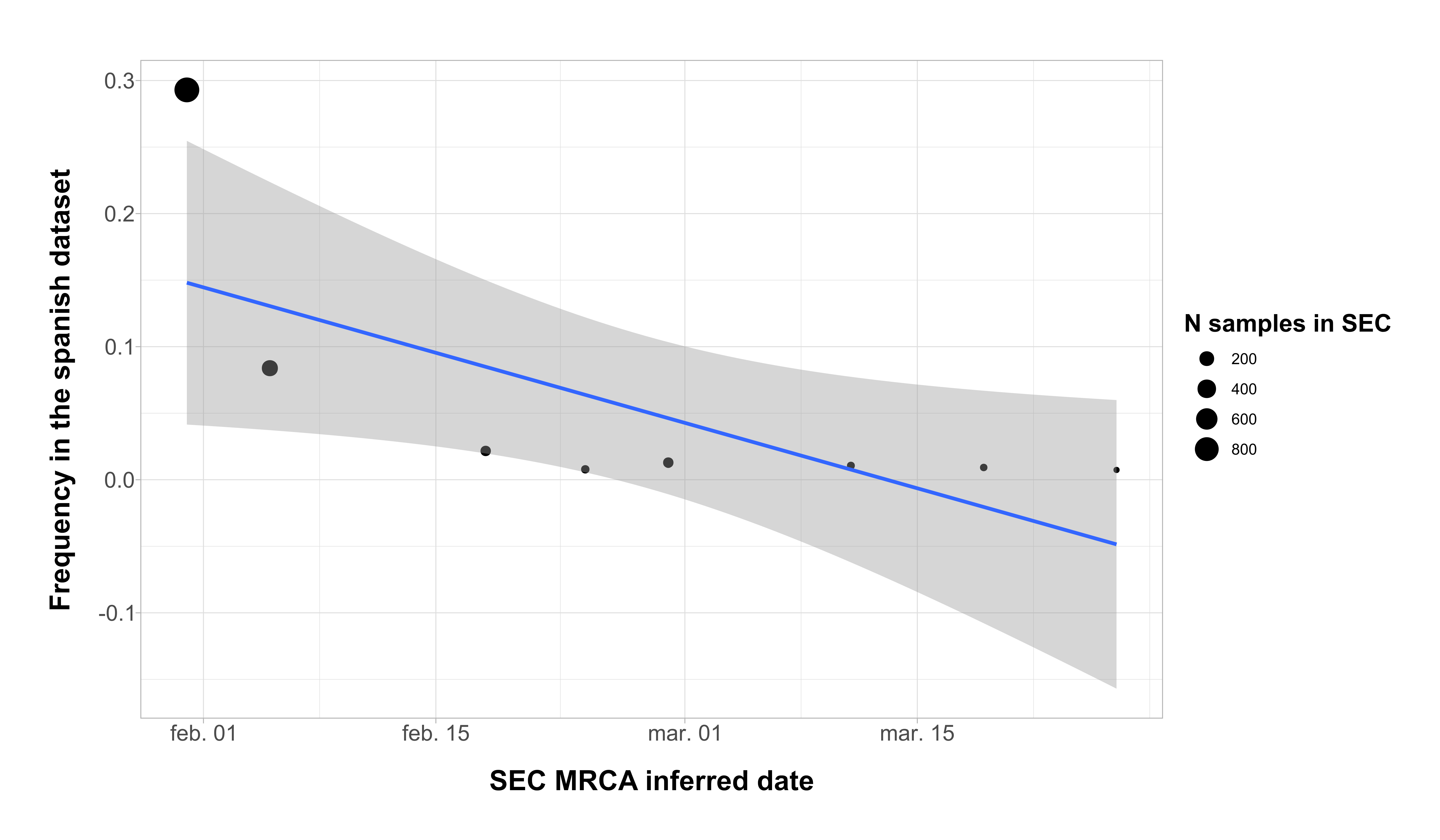

### Figure_S7.png

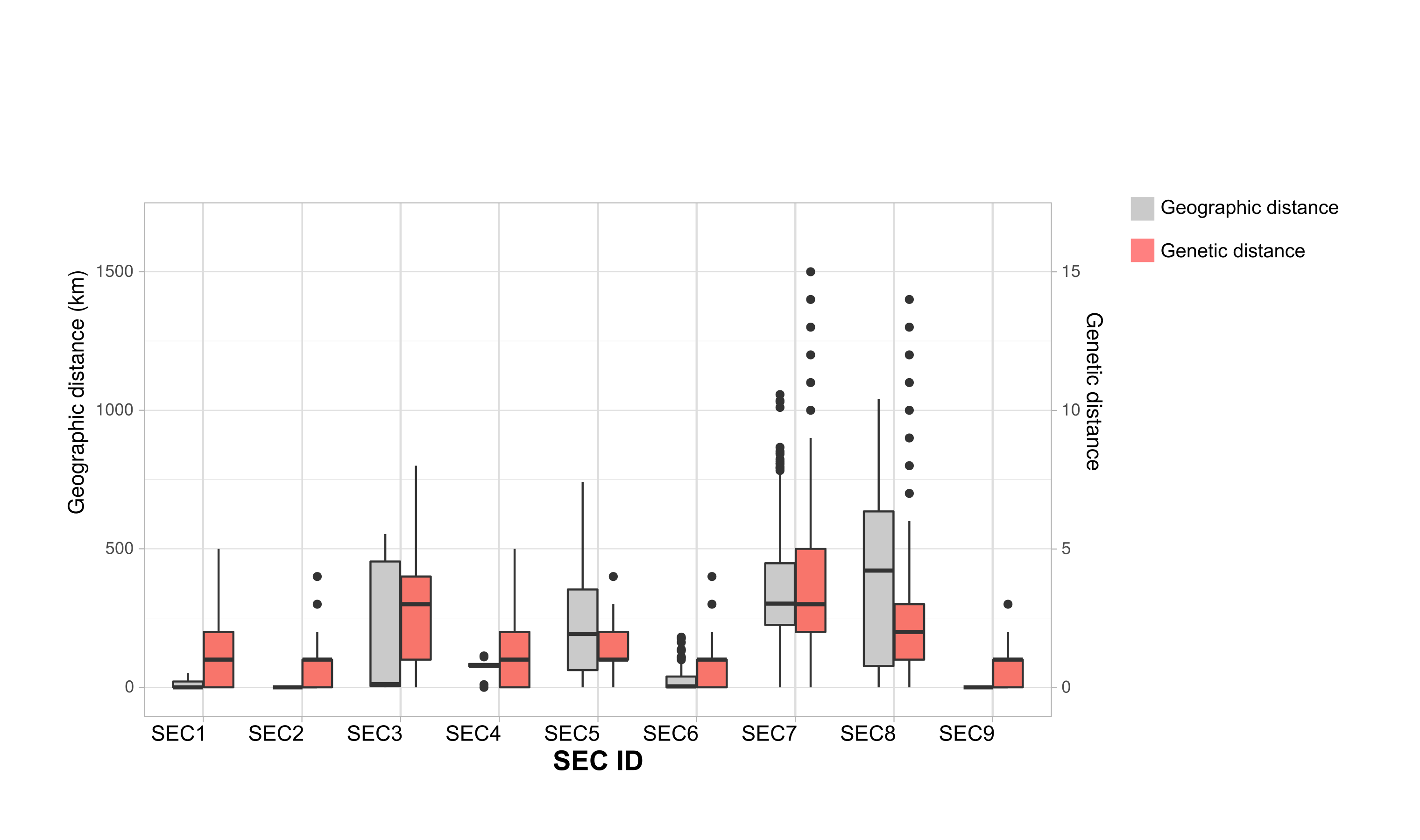

### Figure_S8.png

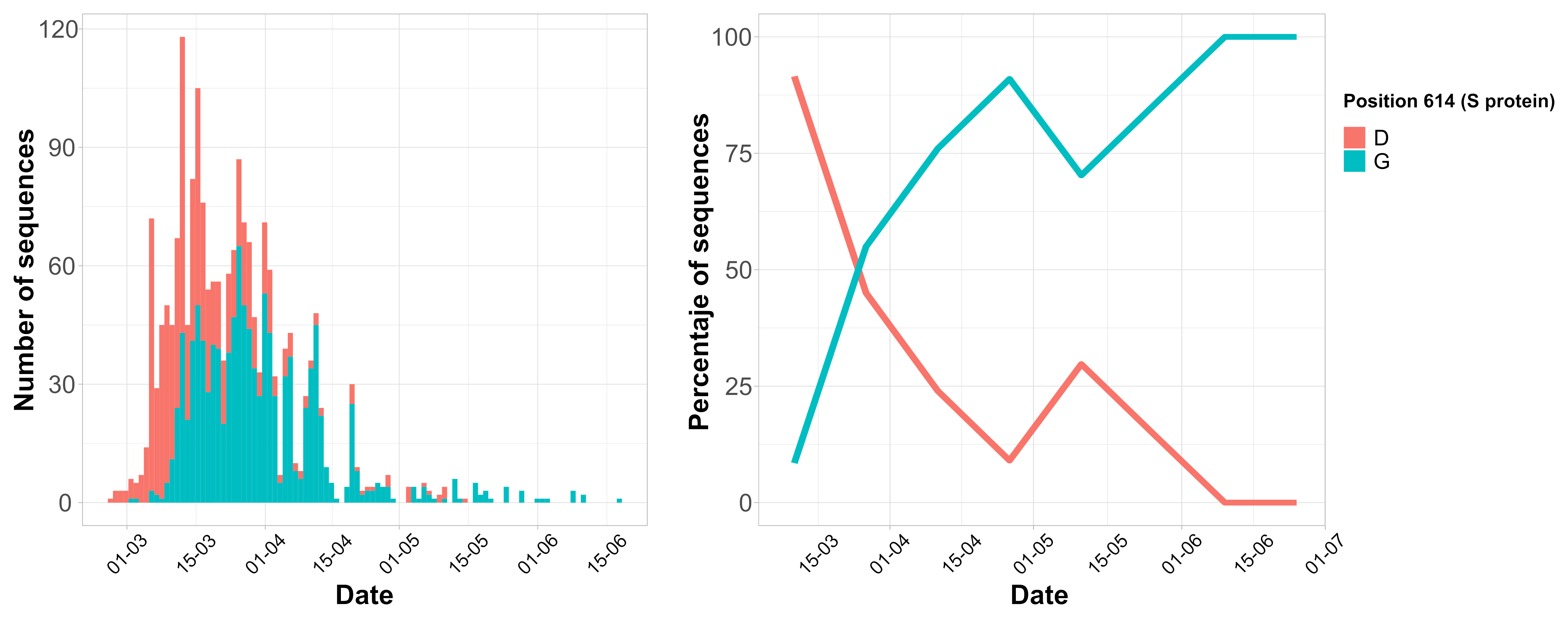

### Figure_S9.png

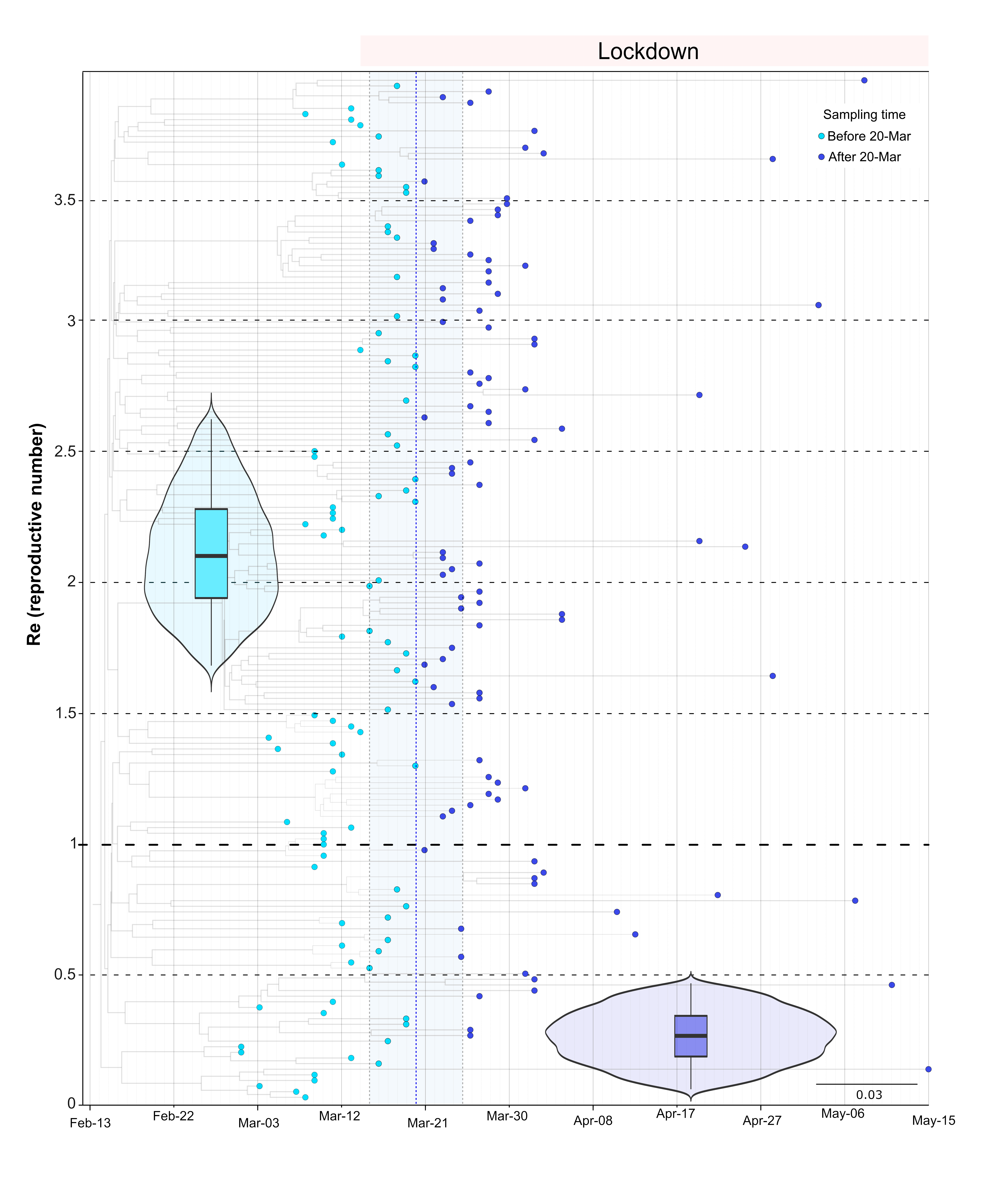

### Figure_S10.png

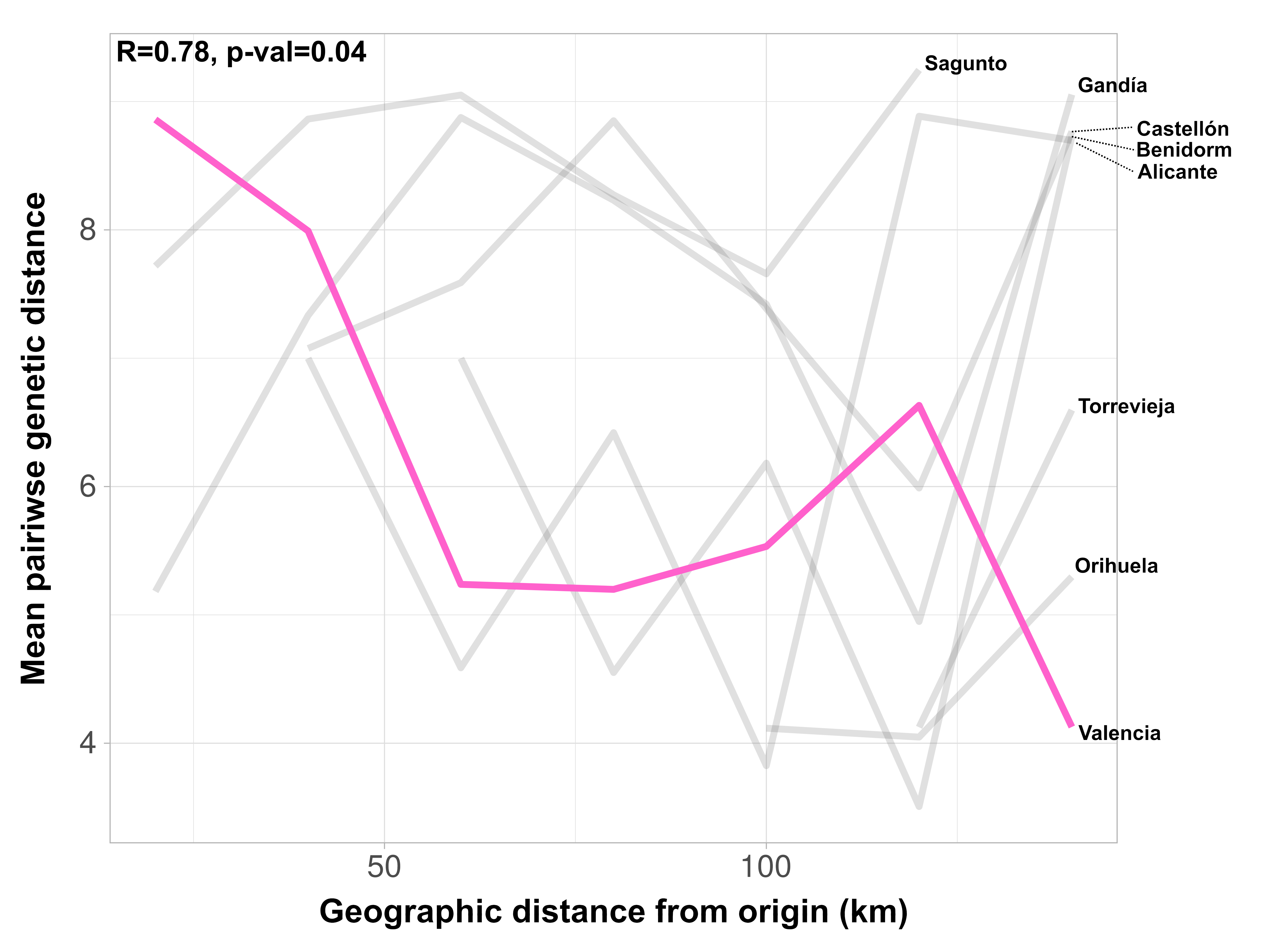

### Figure_S11.png

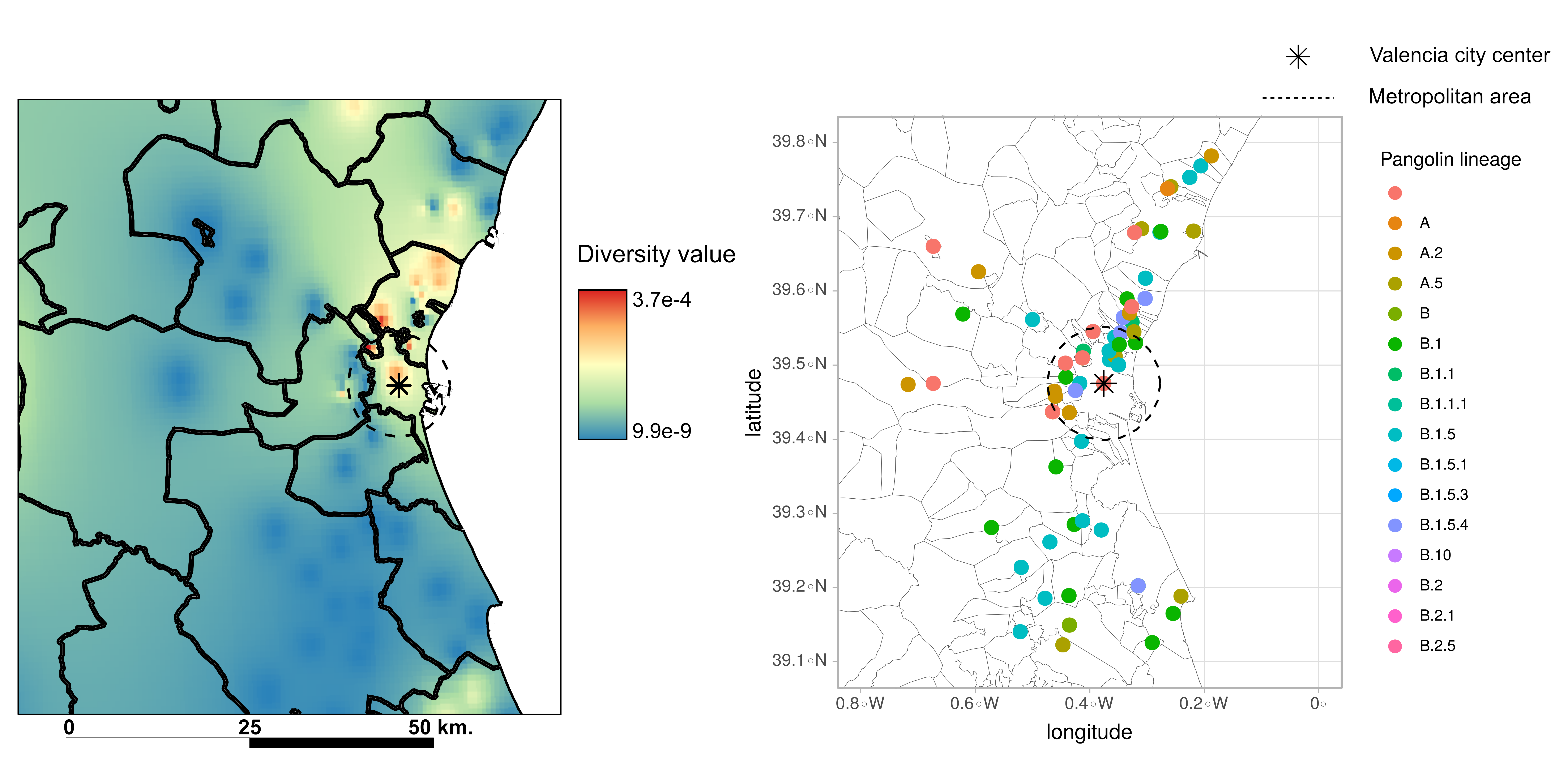
