## Supplementary notes, materials and methods for "The first wave of the Spanish COVID-19 epidemic was associated with early introductions and fast spread of a dominating genetic variant"

#### Large metropolitan areas as sources of viral genetic diversity

We observed a heterogeneous distribution of SARS-CoV-2 genetic diversity across Spain, both at regional and local levels (Figure 1b). Some regions concentrate different viral lineages while others have low genetic diversity and are dominated by one or few lineages. We found a weak correlation between the mean viral genetic diversity and the population density of each

municipality (ρ=0.32, p-value=0.01), suggesting that more densely populated areas exhibit higher diversity of SARS-CoV-2 strains compared to low-density regions.
We hypothesize that large cities acted as sources of viral genetic diversity during the pandemic, with their “economic-influence” areas (e.g. urban outdoor areas) acting as sinks. To test this, we used the autonomous region of Comunidad Valenciana as a case study, since this was the region for which we have the largest sampling. We calculated the genetic diversity and the mean pairwise distances between samples belonging to different municipalities and correlated these values against their geographic distance to the regional capital city, Valencia. Results (Figure S10, Figure S11) suggest that the city of Valencia was the center of the genetic diversity within the region; the genetic diversity decreases with geographic distance to Valencia up 70-80km away from the city center. This result agrees with a “model of isolation” by distance, where only part of the genetic diversity of the original population (Valencia) spreads to the surrounding areas; it is also compatible with patterns of population mobility within large cities before the installment of a national lockdown.

#### Spread of SECs across the country

We observed a correlation between the time of introduction and SEC size (Figure S6). In the beginning, before the adoption of non-pharmaceutical interventions, strains were transmitted without control and generated larger clusters. After restrictions were implemented, transmission was controlled and smaller clusters were observed. Regardless of their introduction time, some strains could spread locally while others dispersed throughout the rest of Spain, generating different dispersion patterns. To study this, we first analyzed the geographic distribution of samples within each SEC (Figure 2d). We noticed that the largest and oldest SECs (SEC7 and SEC8) were the most widely distributed and were present in 10 or more Spanish regions. Conversely, the smallest and youngest SECs (SEC1 and SEC2) spread only locally, in Andalucía and País Vasco respectively, in agreement with its MRCA ancestor occurring after the lockdown. The remaining SECs displayed varying but generally narrower dispersal patterns compared to SEC7 and SEC8, and are present in 2 up to 6 Spanish regions. We also analyzed the distribution of geographic distances (kilometers) between samples within the same SEC. An ANOVA analysis (adjusted p-value << 0.01) divides SECs into three different categories: SECs 1,2,4,6 and 9 showed very limited geographical dispersion; SEC3 and 5 showed medium values and SEC7 and 8 exhibited the widest geographical distribution. The smallest SECs include samples within a range of about 0-58 km (Figure S7), whereas the mean distance between samples in SEC7 is 323 km (interquartile range 225-447 km) and 371 km (interquartile range 76-634 km) in SEC8. This supports the hypothesis that SECs 7 and 8 spread rapidly throughout the country after their first introduction. Interestingly, despite their wider geographic range compared to the rest of SECs, they do not exhibit a larger accumulation of mutations (Figure S7).

#### Impact of lockdown

In order to evaluate the effectiveness of the restriction movements on the spread of the pandemic in Spain, we performed a Bayesian birth-death skyline (BDSKY) analysis in order to estimate the timing and magnitude of changes in the effective reproductive number ($R_e$), as a measure of the average secondary cases generated by an infected person, for the most successful SECs. For SEC7 the results suggested a strong evidence for a change in $R_e$ around 20th March with a decrease below one, after the restrictions implemented by the Spanish Government on 14th March(Figure S9).

In the case of the SEC8, the decrease of the $R_e$ was estimated after 9th March, a bit earlier than the lockdown implementation, this is probably due to the bias produced by the multiple introductions that cannot be differentiated from transmission events by the birth-death skyline model as can be observed in the multiple and basal tree bifurcations (Figure 3c).

Doubling time, as measure of the amount of time in which the incidence doubles [1], was also calculated for both SECs as 365 days/year × ln(2) / ($R_e$ × δ)- δ), where δ is known as the become uninfectious rate and is the inverse of the duration of infection, which we assume to be 10 days, or 10/365 years (such that δ=36.5 $year^{-1}$). Before the corresponding $R_e$ changing date, the doubling time was estimated at 6.3 days (95% HPD: 4.3-10.2 days) for SEC7 and 3.3 days (95% HPD: 2.7 - 4.1 days) for SEC8. After changing date $R_e$ value is below 1, the number of cases is decreasing, such that the absolute value of the doubling time is in fact the halving time of the number of infections, i.e the time required for the number of cases to halve. The post-peak halving times are approximately 9.5 and 9.0 days for SEC7 and SEC8 respectively, reinforcing the results of a pandemic reduction after the confinement.

### SUPPLEMENTARY MATERIALS AND METHODS

#### SeqCOVID sampling and sequencing

RNA samples were received from different hospitals, and confirmed as SARS-Cov-2-positive by RT-PCR by Microbiological Services. Samples were the remaining RNA extracts from naso- and oropharyngeal clinical specimens employed for diagnosis. The use of such samples have been approved by the ethics committee Comité Ético de Investigación de Salud Pública y Centro Superior de Investigación en Salud Pública (CEI DGSP-CSISP) Nº 20200414/05. RNA was retro-transcribed into cDNA and SARS-CoV-2 complete genome amplification was conducted in two multiplex PCR, accordingly to openly available protocol developed by the ARTIC network [2] using the V3 multiplex primers scheme [3]. Two resulting amplicon pools were combined and used for library preparation. Genomic libraries were constructed with the Nextera DNA Flex Sample Preparation kit (Illumina Inc., San Diego, CA) according to the manufacturer's protocol, with 5 cycles for indexing PCR. Whole genome sequencing was carried out in the MiSeq platform (2×200 cycles paired-end run; Illumina).

The sequences obtained went through a bioinformatic pipeline based on IVAR [4], which is open source and can be accessed at https://gitlab.com/fisabio-ngs/sars-cov2-mapping. In short, the pipeline goes through the following steps: 1) Removal of the human reads with Kraken [5]; 2) filtering of the fastq files using fastp v 0.20.1 [6] (arguments: --cut tail, --cut-window-size, --cut-mean-quality , -max_len1 ,-max_len2 ); 3) mapping and variant calling using IVAR v 1.2; 4) quality control assessment with MultiQC [7].

#### Global alignment and phylogenetic reconstruction

To build the global alignment, we downloaded and concatenated all non-spanish sequences present in GISAID [8] on 21st June that passed strict filtering criteria: i) sequences should have more than 29,000 bp length, ii) verified insertions/deletions, iii) less than 1% of Ns and less than 0.05% of unique amino acid mutations (compared with other sequences in GISAID).

Later, we added all Spanish sequences deposited in GISAID up to July 29th. The final alignment constructed included 32,914 sequences. The accession numbers of the sequences used in this study can be found in Table S1.

Sequences were aligned against the SARS-CoV-2 reference genome [9] using MAFFT [10]. Specific positions that have been reported to be problematic for phylogenetic reconstruction [11] were masked, following the procedure described by Rob Lanfear [12], using the mask_alignment.sh script.

Finally, a maximum-likelihood (ML) phylogeny was reconstructed using IQTREE [13] with the GTR model and based on the complete masked genome alignment. This phylogeny was rooted to the SARS-CoV-2 sequence obtained in Wuhan on 24/12/2019 (GISAID ID: EPI_ISL_402123).

#### Identification of introductions and transmission clusters

We identified transmission groups between Spanish sequences by inspecting the global phylogeny (32,914 leaves) and searching for Spanish sequences (or groups of) that were embedded within sequences with other geographic origins. Given the general low diversity among sequences, most phylogenetic nodes ended up being polytomic in the maximum-likelihood tree. Because of this, we defined three different transmission scenarios: i) strains that represent introductions in Spain but differ from those from other countries and form well defined transmission groups (‘candidate transmission clusters’); ii) strains introduced into Spain that are equal to other Spanish sequences and that are also equal to sequences from other countries (‘zero distance clusters’); and iii) Spanish sequences found within groups of sequences from other countries and which are not phylogenetically near any other Spanish sequences (‘unique’). The ‘candidate transmission clusters’ were identified as monophyletic groups of sequences composed exclusively by Spanish sequences in the phylogeny. The ‘zero distance clusters’ were identified as Spanish sequences that share a common ancestor and that are at 0 SNP distance from each other. Finally, The ‘unique’ sequences were identified as those sequences which do not share their most recent ancestor with any other Spanish sequence.

Next, we inferred how many of these transmission groups have a potential contagion date for their first case that predates the start of mobility restrictions, on 14th March, by subtracting 14 days to the diagnosis date.

Finally, we wanted to investigate the international origin of these introductions. For each of the identified groups or ‘unique’ sequences with an inferred contagion date before 14th March, we looked for the closest non-Spanish sequence in the phylogeny with a diagnosis date predating the first case of the transmission group. As the current consensus is that the pandemic began in Asia and later it moved to Europe, we considered only those sequences with an Asian or European origin as potential sources of introductions.

#### SEC alignment and phylogeny

Using the global phylogeny, we identified nodes which had at least 20 leaves and in which at least 50% of these correspond to Spanish sequences. Next, for each of these nodes or clades we reconstructed an alignment of the complete masked genomes including:

a. The sequences that belong to the identified clade
b. 11 basal sequences from Wuhan acting as an 'anchor' for the phylogeny (Table S1).
c. A subset of 51 representative sequences, each one from a different pangolin lineage, selected to maximize the global SARS-CoV-2 genetic diversity (Table S1, downloaded from GISAID on 2020-07-20).

For each of these alignments we inferred a ML phylogeny, using IQTREE [13], with the model GTR, 1,000 fast-bootstrap replicates and rooted to the Wuhan sequence (EPI_ISL_402123). Then, in the resulting phylogeny we identified less inclusive nodes embedded within the above identified clades and had a bootstrap support value > 80. These clades were named as potential Spanish epidemic clades (SEC). The iTOL tool [14] was used for phylogenetic visualization.

#### SEC8 detailed analysis

To get more detail on SEC8 phylogenetic structure, and to evaluate if mobility restrictions were effective to hinder SEC8 transmission, we enriched the original SEC8 phylogenetic tree with all the isolates of this clade sampled from February to October by the SeqCOVID consortium (959 sequences in total). Later, epidemiological information was included and plotted in the tree using the iTOL tool [14].

SEC8 potential superspreading events were defined as groups of more than 10 sequences, having at least 1 SNP in common and having a within sequence median distance from 1 to 3 SNPs.

#### Population genetics and differentiation geography

Geographic distance between sequences were computed using the GPS coordinates of the patient residence city and applying the Vicenty (ellipsoid) method. Genetic diversity was calculated with two different methods: i) genetic distance between each pair of samples in number of substitutions (SNPs), and ii) number of base substitutions per site averaged over all sequence pairs in a group of sequences. Both values have been estimated using the MEGA software [15], skipping one position when a gap is found in the two compared sequences.
Demographic data for all Spanish regions and municipalities were downloaded from INE (https://www.ine.es/), and had been updated on 1$^{st}$ January, 2020.

The genetic diversity heatmap of the Comunidad Valenciana autonomous was generated with QGIS v.3.14.16-Pi [16], using the inverse distance weighting (IDW) algorithm to interpolate the mean genetic diversity of each municipality for which we had at least two sequences.

To compare the genetic and geographic distance distribution between the different SECs, we used a one-way ANOVA test, followed by multiple pairwise-comparisons of the between-groups mean with a Tukey HSD test.

#### Dating analyses

To estimate the most recent common ancestor (MRCA) of each of the nine SECs defined above, a multi-sequence alignment was performed including the 11 samples belonging to basal phylogenetic clades and the 51 representative sequences from different lineages (Table S1). Before phylogenetic dating, root-to-tip regression of genetic divergence against sampling dates was performed to investigate the molecular clock signal of SECs using TempEst v 1.5.3 [17]. We implemented a coalescent Bayesian exponential growth model available in Beast 2.6 [18] with the HKY+Γ model of nucleotide substitution. Tree priors were defined as follows: for effective population size we used a lognormal distribution (M=1, S=2) and for growth rate a Laplace distribution (M=0, S=100). The uncorrelated lognormal relaxed clock was selected as the best fitting clock model using Bayes Factor comparisons of strict and relaxed clocks based on path sampling/stepping stone analysis [19]. Clock priors were defined as: ucld.mean: lognormal distribution with mean in real space = $1.4 \times 10^{-3}$ subs/site/year and ucld.stdev = $5 \times 10^{-2}$. Parameters were estimated using Markov Chain Monte Carlo (MCMC) Bayesian inference, with $5 \times 10^{7}$ steps-long chains with exception of SEC7 and SEC8, for which longer chains were run ($1 \times 10^{8}$), in all cases a total of $10^{5}$ steps were sampled in the log files. For all analysis, three independent runs starting from different seeds were conducted in order to ensure convergence, then combined with LogCombiner v 2.6.3 after removing the initial 10% of the MCMC as burn-in. Adequate mixing of parameters and convergence among runs were assessed using Tracer v 1.7.1 [20] by verifying that each parameter reached an effective sampling size (ESS) above 200 and that traces showed stationarity and good mixing. The final posterior distribution contained a total of 9000 trees, annotated with Treeannotator v 2.6.3 and visualized in FigTree v 1.4.3 [21]

#### Phylodynamics analysis to estimate $R_e$

To estimate discrete changes in $R_e$ through for the two largest epidemic clades SEC7 and SEC8. We used a Bayesian birth-death skyline model (BDSKY) with serial sampling [22] implemented in BEAST v 2.6 [18]. BDSKY uses an episodic, piecewise birth-death model in which the parameter is allowed to change at discrete points in time, with the magnitude and timing of changes estimated from the data. In our analysis, we set two intervals wherein $R_e$ is constant and estimated the date with most evidence for a change in $R_e$. To this end, we set a uniform prior distribution. $R_e$ was estimated before and after the changing time. Same parameters as above were used but fixing the clock rate and the become uninfectious rate ($\delta$ = 36.5 years$^{-1}$) in accordance with consistent global estimates of an infectious period of 10 days [23]. In order to avoid bias in the model parameters due to constant sampling proportion assumed by BDSKY models, this parameter was set to zero before the first sample date using TreeSlicer (https://github.com/laduplessis/skylinetools/wiki). For this analysis a 1 x $10^7$ and 4 x $10^7$ steps-long chains were used for SEC7 and SEC8 respectively. Results were inspected with Tracer (v 1.7.1) [20] by verifying that every parameter had effective sampling sizes above 200 and well mixing was obtained. Doubling time was calculated from the parameters estimated by BDSKY model in which growth rate(r) = $(R_e * \delta) - \delta$ and doubling time = $\ln(2) / r$.

#### Statistical analysis

All statistical analyses were carried out using the R statistical language [24]. Packages ape [25], treeio [25,26], doParallel [27] and foreach [28] were used for phylogenetic manipulation and analysis. We additionally used packages geosphere [29] , lwgeom [30], sp [31], sf [32] and rgeos [33] to calculate the geographic distances between samples and the geographical representation in the data. The ggplot2 R package [34] was extensively used for analysis and data plotting.

### Data availability

The analysis pipeline used to map and analyze the sequences is available at https://gitlab.com/fisabio-ngs/sars-cov2-mapping. All the genomic sequences used in the analyses are available in the GISAID database, and the accesión numbers can be found in Table S1.
